# Covid-19 and Population Age Structure

**DOI:** 10.1101/2020.05.31.20118349

**Authors:** Ajit Haridas, Gangan Prathap

## Abstract

Epidemiological studies suggest that age distribution of a population has a non-trivial effect on how morbidity rates, mortality rates and case fatality rates (CFR) vary when there is an epidemic or pandemic. We look at the empirical evidence from a large cohort of countries to see the sensitivity of Covid-19 data to their respective median ages. The insights that emerge could be used to control for age structure effects while investigating other factors like cross-protection, comorbidities, etc.

## Introduction

Two recent studies (Haridas & Prathap 2020a,b) presented a relation between the deaths from Covid-19 and influenza for many countries. One of the confounding factors which had to be eliminated was that of population age structure. Epidemiological studies, for e.g., Docherty et al. (2020), suggest that age distribution of a population has a non-trivial effect on how morbidity rates, mortality rates and case fatality rates (CFR) vary when there is an epidemic or pandemic. Dudel et al. (2020) examined the age specific CFR in 7 countries and found that it accounts for a substantial fraction of the variation between the countries in their sample study. Dowda et al., (2020) suggests that young age structure of African countries is protective of severe infection and thus the lower than expected number of cases detected in Africa (despite extensive trade and travel links with China). Here we report the effort we made to collect empirical evidence from a large cohort of countries to see the sensitivity of Covid-19 data to their respective median ages. The insights that emerged was used to control for age structure effects while investigating other factors like cross-protection, comorbidities, etc. as in Haridas and Prathap (2020a,b).

## A suitable age structure variable

It is clear that one needs an appropriate age structure variable. From https://databank.worldbank.org/source/world-development-indicators# we can collect the population for three age groups as a % of total population: ages 0–14, ages 15–64 and ages 65 and above for more than 250 countries for 2018 (last updated on 09-04-2020 when last accessed). We also collect from https://en.wikipedia.org/wiki/List_of_countries_by_median_age the median ages for a comparable number of countries. In economic studies a ‘dependency ratio’, defined as the number of children and retired persons to those of working age (Leff 1969) is used (here this can be defined as (15–64)/(00-14 + 65UP). We performed many correlation studies with candidate age structure variables (not reported here) and found that the median age is a reliable measure of age structure than dependency ratio for the present purpose. We shall use it in what follows.

## Methodology

As done earlier in Haridas & Prathap (2020a, 2020b)), we obtain the actual or estimated count of cases and deaths from Covid-19 as of 30/04/2020 for many countries from https://www.worldometers.info/coronavirus/. The case fatality rate (CFR) for a country was defined as the ratio of cumulative cases and cumulative deaths. We plot cases per million, deaths per million and CFR against median age for 156 countries for which complete data was available. As the cases, deaths and CFRs range over several orders of magnitude, and median age ranges from 15.4 to 47.3, semi-log plots were the best choice.

## Results and discussions

Figure 1 shows the scatter plot of Covid-19 cases per million of population vs the median age on semi-log scales. From “young” to “old” nations, there is a clear stratification seen from low values of morbidity rates to values which are orders of magnitude higher. We have Niger (NER), Equatorial Guinea (GNQ), Djibouti (DJI), Qatar (QAT), Iceland (ISL) and Luxembourg (LUX) on the high end. Iceland is here because it has the largest number of tests per million so far and so reports a very large number of cases most of which may be asymptomatic. The very rich (Qatar and Luxembourg) and the very poor (Niger, Equatorial Guinea and Djibouti) are on the same high cases per million boundary. In a similar manner, young and old and rich and poor, like Yemen (YEM), Vietnam (VNM) and Japan (JPN) show the low boundary limits. Note that there no zero case cases here!

**Figure 1.**
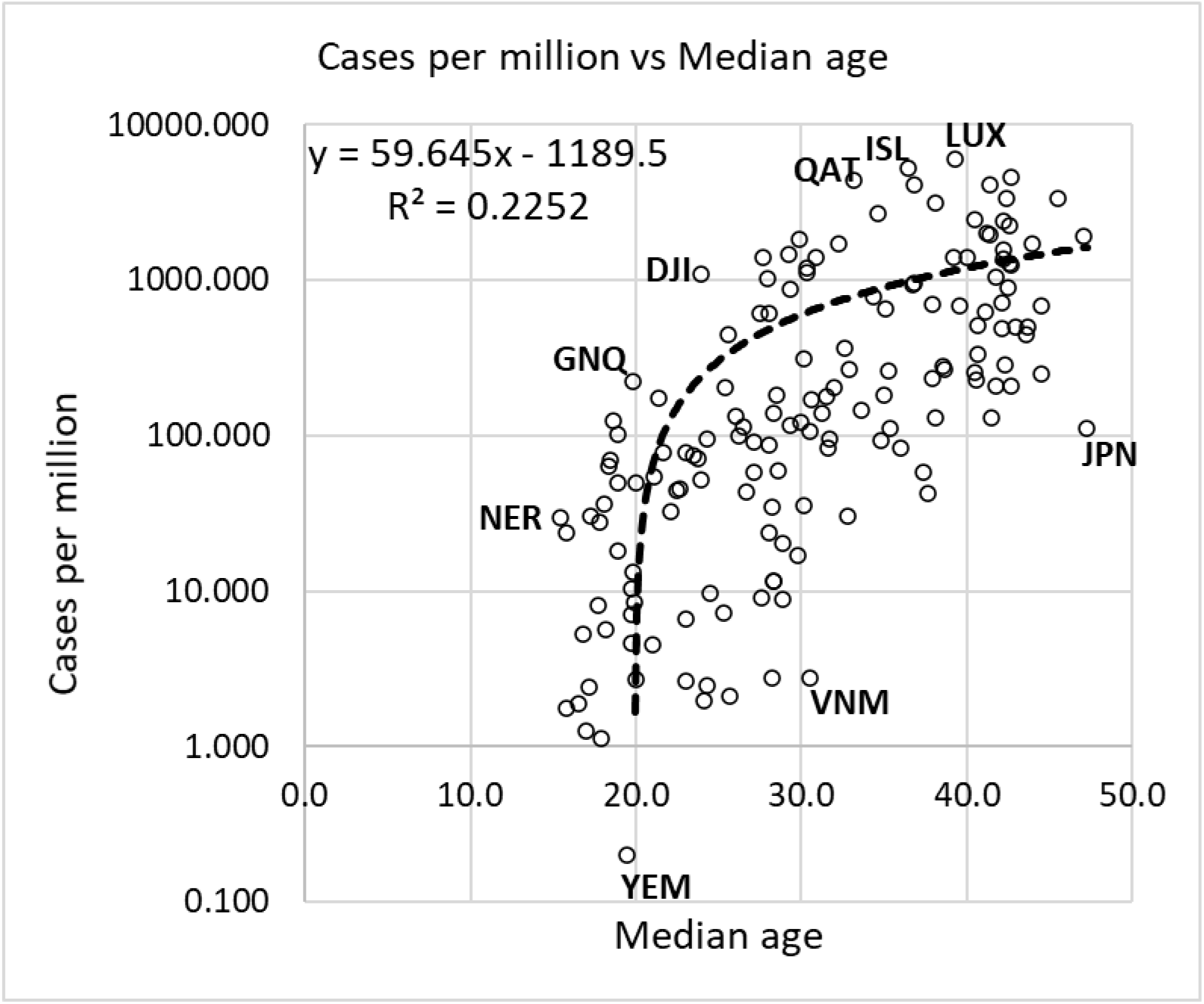
Scatter plot of Covid-19 cases per million of population vs the median age.

Figure 2 shows the scatter plot of Covid-19 deaths per million of population vs the median age on semi-log scales. Unlike the previous case, here there are several zero death cases at the time of collecting the data and these cannot be plotted on log scales. Again, from “young” to “old” nations, there is a clear stratification seen from low values of mortality rates to values which are orders of magnitude higher. We have Niger (NER), Liberia (LBR), Ecuador (ECU), and Belgium (BEL) on the high end. In a similar manner, young and old and rich and poor, like Ethiopia (ETH), Myanmar (VNM) and Japan (JPN) appear on the low non-zero boundary limits.

**Figure 2.**
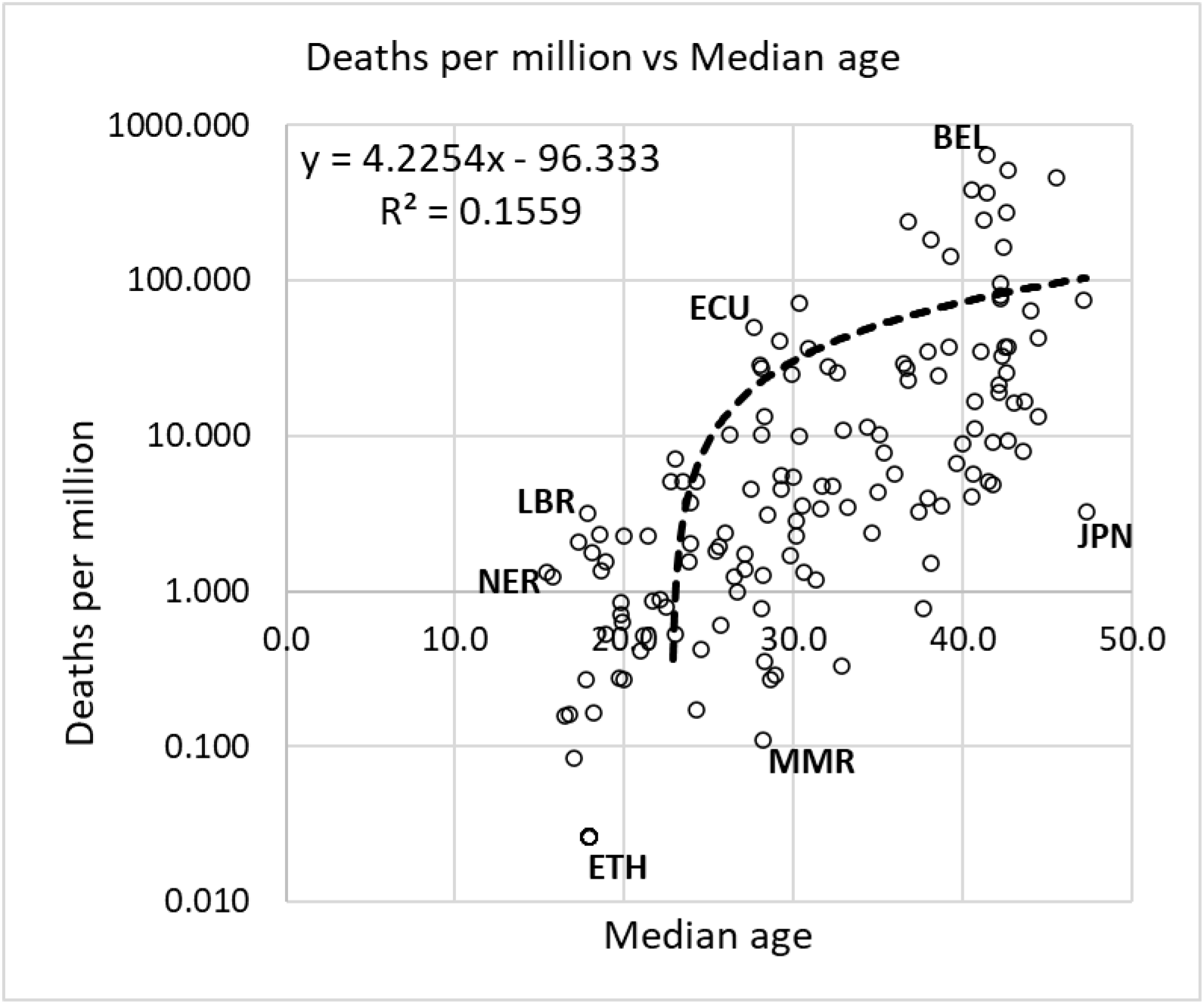
Scatter plot of Covid-19 deaths per million of population vs the median age.

In both Figs. 1 and 2, as a semi-log representation is used, we expect an exponential fit which will then be captured by the linear trendline using the feature already available in Excel. We see that as a general rule, older populations show higher morbidity and mortality rates, as is to be expected for Covid-19. The linear trend line, show that the relation between median age and cases or between median age and deaths is not linear.

Figure 3 shows the scatter plot of Covid-19 CFR vs the median age on semi-log scales. Again, there are several zero death cases at the time of collecting the data and these cannot be plotted on log scales. But now, from “young” to “old” nations, although there is a clear stratification seen from low values of CFR to values which are orders of magnitude higher, we find that in general, there is no sensitivity to median age. That is, the exponential fit which is anticipated by the linear trendline using the feature already available in Excel shows that there is very little variation, As a general rule, older populations do not show higher CFR, as is to be expected for Covid-19. There are other confounding factors involved. It is interesting to note that Qatar (QAT) and Singapore (SGP) are egregiously placed as outliers with significantly lower CFR; both countries have a very large young migrant population.

**Figure 3.**
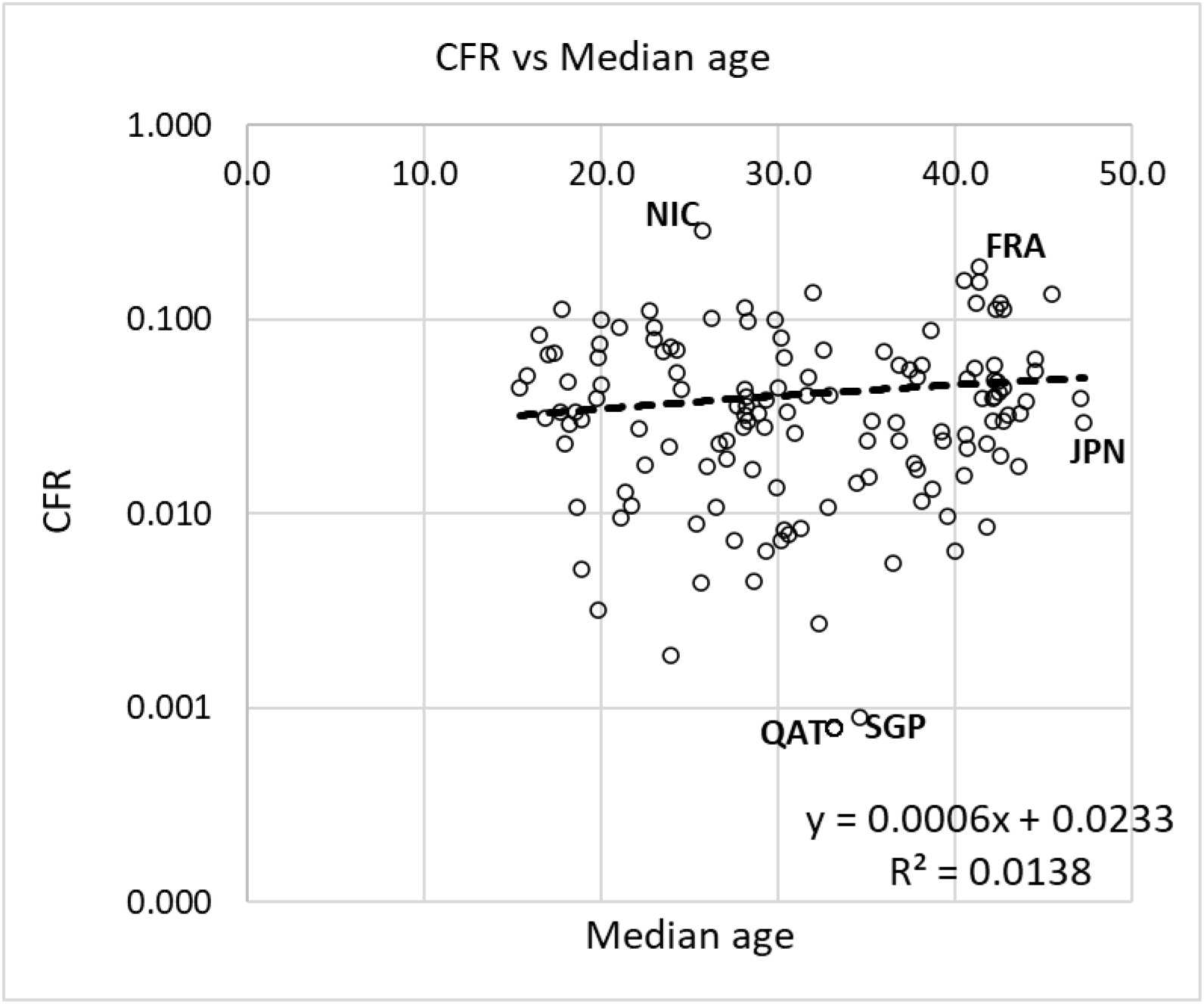
Scatter plot of Covid-19 CFR vs the median age.

Clearly other confounding factors contribute in the country-wise variability in CFR within a median age band. However, as it is known that older individual have greater mortality risks to Covid-19, the poor sensitivity of CFR to country median age is further examined. We carry out a filtering process to extract any underlying trend. The country-wise data on cases was sorted by median age and the moving average cases per million of cohort of 17 countries (17 countries correspond to median ages in 15 – 20 year bracket) was plotted against median age (Figure 4). Two distinct trends are seen in Figure 4 – A nonlinear increase in cases per million up to 35 year median age. Thereafter, cases per million are nearly independent of age, except for a slow declining trend with age.

**Figure 4.**
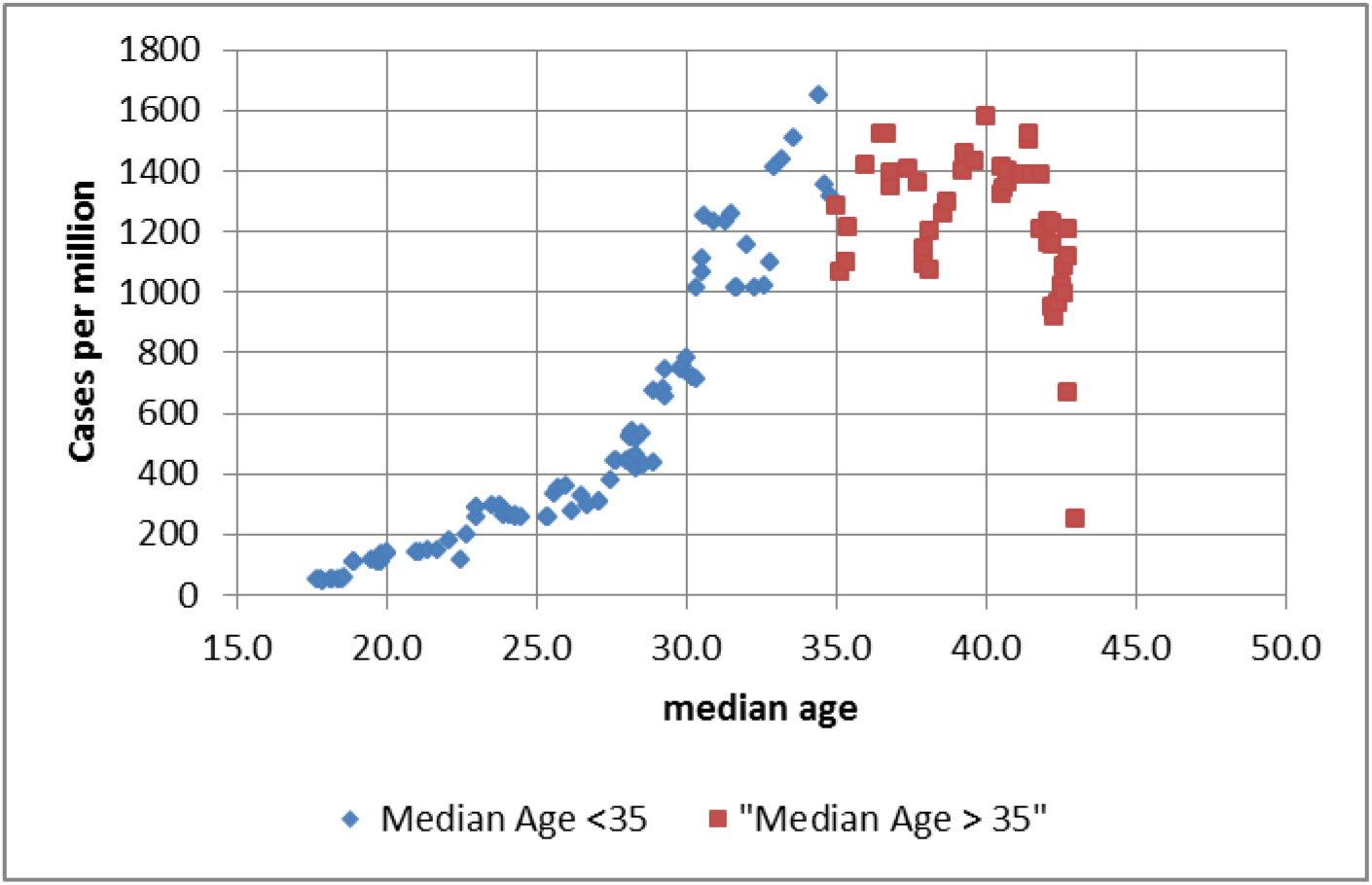
The cases per million averaged across 17 median age cohort countries is plotted against median age.

Figure 5 shows the deaths per million averaged over the same 17 country median age cohort, plotted against the median age. The trend is similar to Figure 4. Deaths increase with age at greater than linear rate. Above, median age 40, there is no correlation with age, and the trend shows a decline. The variability in deaths per million in the averaging samples above median age 40, is very large as shown in Figure 6. Therefore, the negative trend may be treated with caution.

**Figure 5.**
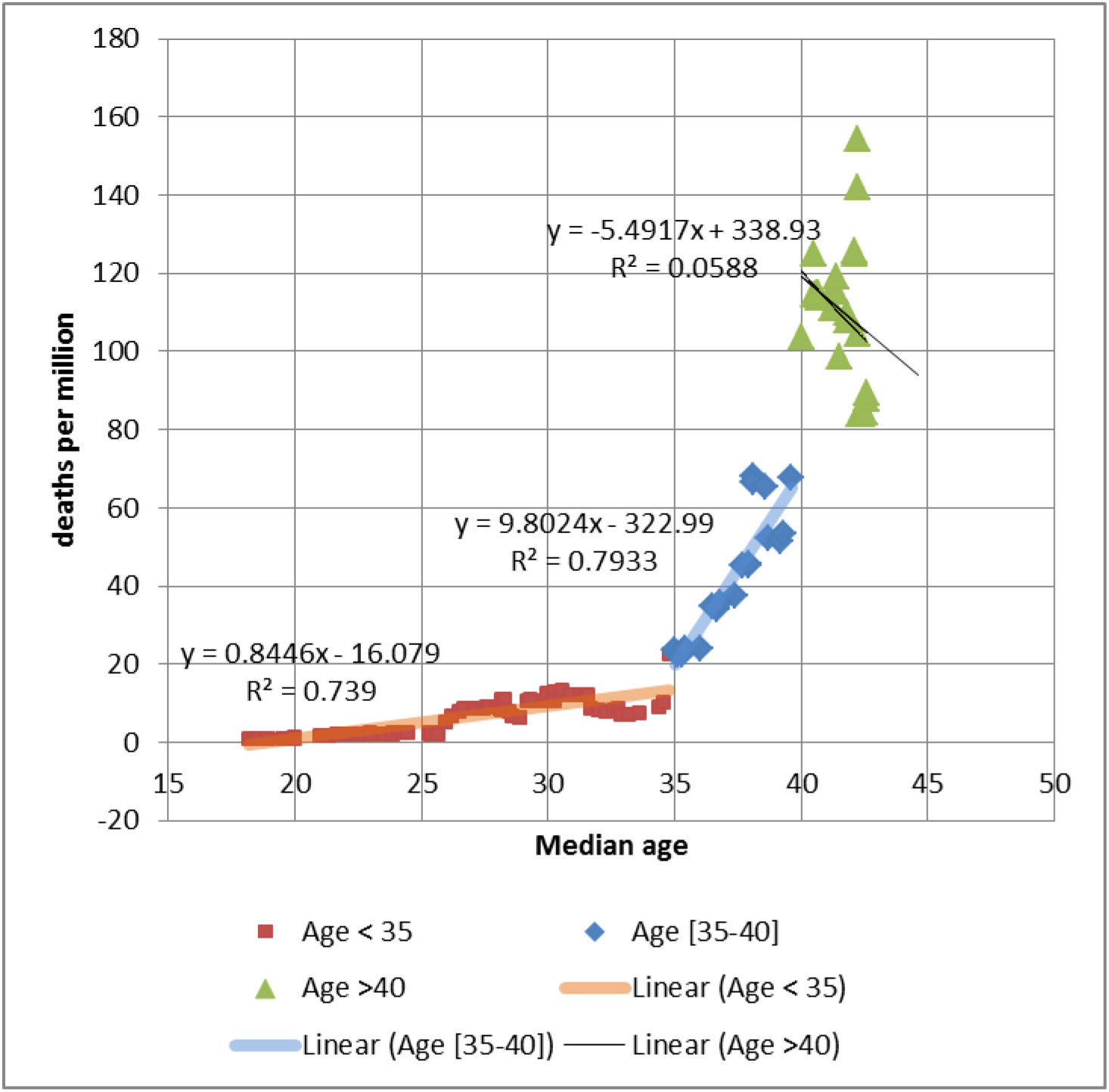
The deaths per million averaged over median ages is plotted against median age.

**Figure 6.**
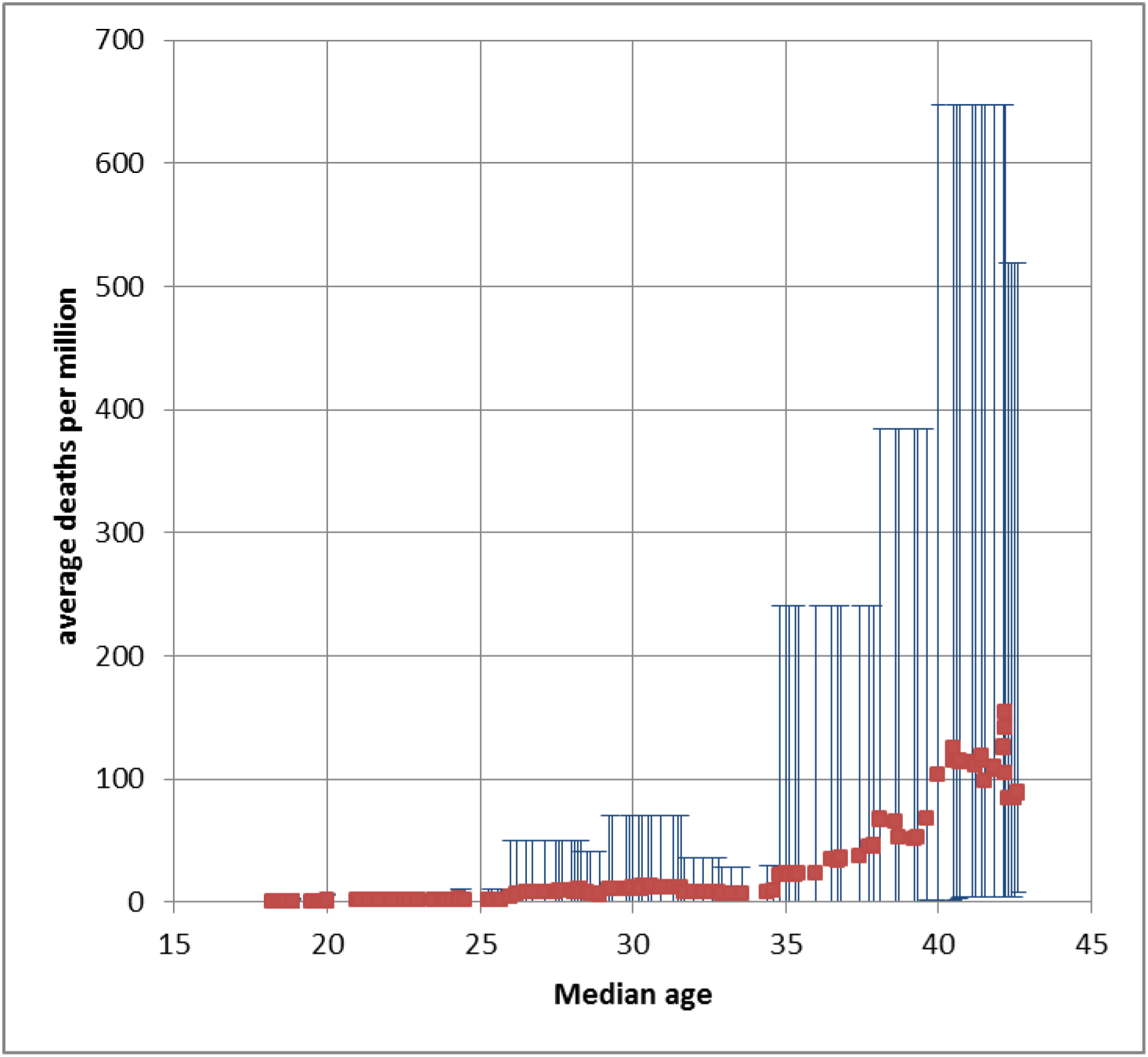
The maximum and minimum within the averaging interval (17) of Figure 5 is plotted along with the average deaths per million against median age.

The filtering procedure on worldwide country-wise data shows a pattern expected from the biology of immune system. We are able to see in the trend an adaptive response that declines with age, and an accumulated memory response, that improves with age. It is remarkable that the variation expected in immunity with the age of an individual is recovered from global country-wise median-age sorted data, by the extensive filtering process to remove the confounding effects.

## Concluding remarks

Using data available in the public domain we showed how COVID-19 morbidity, mortality and case fatality rates vary with population age structure. There is a huge stratification; even within a median age band, mortality and morbidity rates can vary by orders of magnitude. However, this is not true for case fatality rate. For reasons not clear but which suggests that there is the factor of ‘cross immunity’ or ‘cross protection’ to be considered, that prior infection with one virus affords protection against antigenically related ones (Haridas & Prathap 2020a,b).

Extensive filtering of country data reveals a distinct pattern of increasing infection rate with median age followed by steady rate of infection. The trends at multi-country median age level reveal underlying age-based infection and death risk of individuals.

## Data Availability

All data available from public databases

